# Statistical Analysis Plan for the HOPE Trial: a randomized trial on Hemodynamic Optimization of cerebral Perfusion after Endovascular therapy in patients with acute ischemic stroke

**DOI:** 10.64898/2025.12.16.25342410

**Authors:** Pol Camps-Renom, Marina Guasch-Jiménez, Natàlia Pérez de la Ossa, Judit Solà-Roca, Joan Martí-Fàbregas

## Abstract

Optimal blood pressure (BP) management following endovascular treatment (EVT) for acute ischemic stroke (AIS) secondary to intracranial large vessel occlusion remains unestablished. The randomized HOPE trial (Hemodynamic Optimization of Cerebral Perfusion after Successful Endovascular Therapy in Patients with Acute Ischemic Stroke) (NCT04892511) seeks to determine if a strategy of hemodynamic optimization using different systolic BP targets, tailored to the degree of final recanalization, is superior to standard BP management in improving functional outcomes for these patients. This document outlines the final Statistical Analysis Plan (SAP) for the trial. This plan will be executed after the last follow-up is complete and the dataset has been locked.

## 1. Trial Information

Title: A randomized trial on Hemodynamic Optimization of cerebral Perfusion after Endovascular therapy in patients with acute ischemic stroke (HOPE study)

Eudra-CT number: 2021-000022-10

Last approved protocol version: v*5 Date: 10-JUL-2024*

Promoter Ethics Committee: Hospital de la Santa Creu I Sant Pau

Protocol study code: IIBSP-HOP-2021-01

clinicaltrials.gov ID: NCT04892511

## 2. Introduction

The optimal management of blood pressure (BP) after endovascular treatment (EVT) for acute ischemic stroke (AIS) caused by intracranial large vessel occlusion remains uncertain. Four clinical trials have investigated different blood pressure (BP) targets following successful recanalization, yielding varied and sometimes conflicting results.

The BP-TARGET trial found no significant difference in intracerebral hemorrhage (ICH) rates between an intensive systolic BP target of 100–129 mm Hg and a standard target of 130–185 mm Hg after endovascular therapy (EVT) [1]. Conversely, the ENCHANTED2/MT trial was prematurely terminated due to safety concerns, as the more intensive treatment group (systolic BP <120 mm Hg) showed a greater likelihood of unfavourable functional outcome compared to the less intensive group (140–180 mm Hg) [2]. Similarly, the OPTIMAL-BP trial suggested a potential harmful effect of intensive BP lowering (systolic BP <140 mm Hg) compared to a 140–180 mm Hg target in patients with successful recanalization (mTICI 2b or greater) [3]. Finally, the BEST-II trial, a futility-design randomized clinical trial, compared systolic BP targets of less than 140 or 160 mm Hg against the guideline-recommended target of 180 mm Hg or less; it did not meet pre-specified criteria for futility or harm [4].

Despite these findings, the optimal postprocedural BP management after EVT remains unresolved. The trials differ in crucial design aspects, such as BP goals and treatment duration. Moreover, variations in patient populations—for instance, the proportion of mTICI 2b vs. 3 or the prevalence of intracranial artery disease—may have influenced outcomes. Limitations also hinder the generalization of these results, including a low median percentage of time spent within the assigned systolic BP target range and the practice of grouping patients with mTICI 2b, 2c, and 3 as equally successful recanalizations. Pending further randomized data, the AHA guidelines [5] still recommend maintaining BP below 180/105 mm Hg for at least 24 hours in patients receiving endovascular therapy (EVT).

The Hemodynamic Optimization of Cerebral Perfusion after Successful Endovascular Therapy in Patients with Acute Ischemic Stroke (HOPE) randomized trial seeks to determine if a strategy of hemodynamic optimization, which tailors systolic BP (SBP) targets post-EVT based on the final degree of recanalization, is superior to standard BP management in improving functional outcomes for these patients.

## 3. Trial design

HOPE is an investigator-initiated multicenter clinical trial with randomized allocation, open-label treatment, and blinded endpoint evaluation (PROBE design). Subjects have been equally allocated (1:1) to hemodynamic optimization according to the protocol of the trial versus BP management according to current guidelines after EVT. The treatment protocol included two different targets of SBP depending on the recanalization status after the procedure. The complete trial protocol has been published previously [6].

### Study population

Patients have been screened for eligibility in 12 endovascular-capable stroke centers in Spain considering the following inclusion and exclusion criteria.

### Inclusion Criteria

- Anterior circulation AIS secondary to intracranial LVO of the M1, M2, A1 segments, including tandem occlusions, within 24 h of symptom onset.
- Successful recanalization at the end of EVT as defined by a mTICI score ≥2b.
- Prior mRS score <3.
- Informed consent by the patient or his/her legal representative.

### Exclusion Criteria

- ASPECTS <6.
- Vertebral, basilar artery (BA), A2, P1-2, or M3-4 segment occlusions.
- History of intracerebral hemorrhage (ICH)
- Pregnancy
- Unstable or recent (<3 months) coronary artery disease or congestive heart failure
- Aortic, cervical, or cerebral artery dissection; unruptured aortic/cerebral arterial aneurysm; or known arterio-venous malformation.
- History of ventricular arrhythmias
- Use of monoamine oxidase inhibitors
- Risk of hemodynamic infarction based on the presence of non-revascularized intra/extracranial stenosis

### Recruitment period

- First patient: 14 - JUN - 2021
- Last patient: 28 – SEP – 2025

### Early termination

Due to a lower-than-expected enrollment rate and the cessation of funding, recruitment for the HOPE trial was halted early on October 1st 2025, with a total of 453 patients recruited.

### Last-follow-up

The final patient follow-up for the trial is anticipated in mid-January 2026.

### Randomization

A conditional stratified randomization was performed using an online tool (clinapsis.com, Clinical Epidemiology and Healthcare Services) based on two possible recanalization status (mTICI 2b or mTICI 2c/3) and then subjects were equally allocated (1:1) to hemodynamic optimization versus standard care in each recruiting center. The randomization occurred within 1 h after the last angiographic image series and since that time, local investigators had one additional hour to achieve the study BP target (shown in Fig. 1).

**Fig 1:**
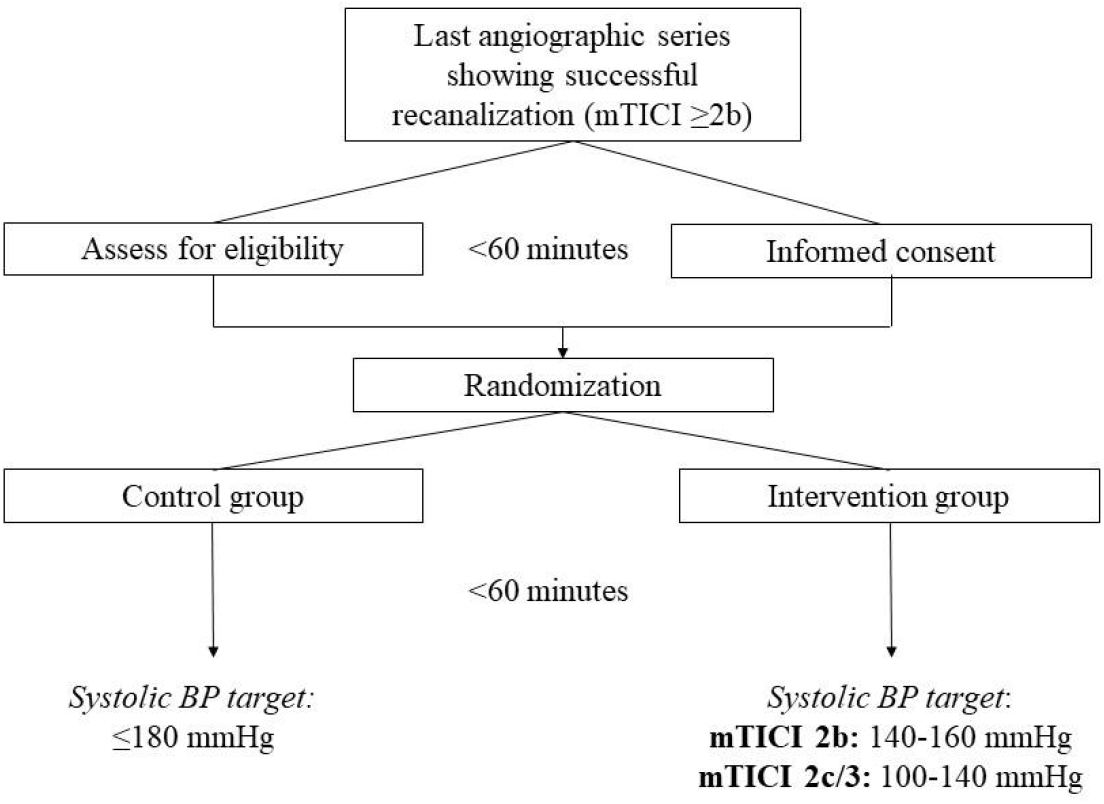
Flow diagram of the study

### Intervention

Patients allocated to the hemodynamic optimization group were assigned to two different SBP targets according to the degree of recanalization after EVT: 100–140 mm Hg for mTICI 2c/3 and 140–160 mm Hg for mTICI 2b. The time interval to reach the desired target was 60 min. Patients allocated to the control arm were treated according to current guidelines (SBP target ≤180 mm Hg) [5]. To achieve the assigned SBP target the study protocol stipulated BP lowering with intravenous labetalol or urapidil, or vasopressor therapy with isotonic saline serum and phenylephrine (shown in Fig. 2 and 3). If the patient achieved the assigned BP target without intervention, no treatment was applied. The study protocol was continued during 72 h after EVT and non-invasive monitorization of BP was performed as follows: 0–24 h, at least every 30 min in the intervention group and at least every 1 h in the control group; 24–72 h, at least every 1 h in the intervention group and at least every 6 h in the control group.

**Fig 2:**
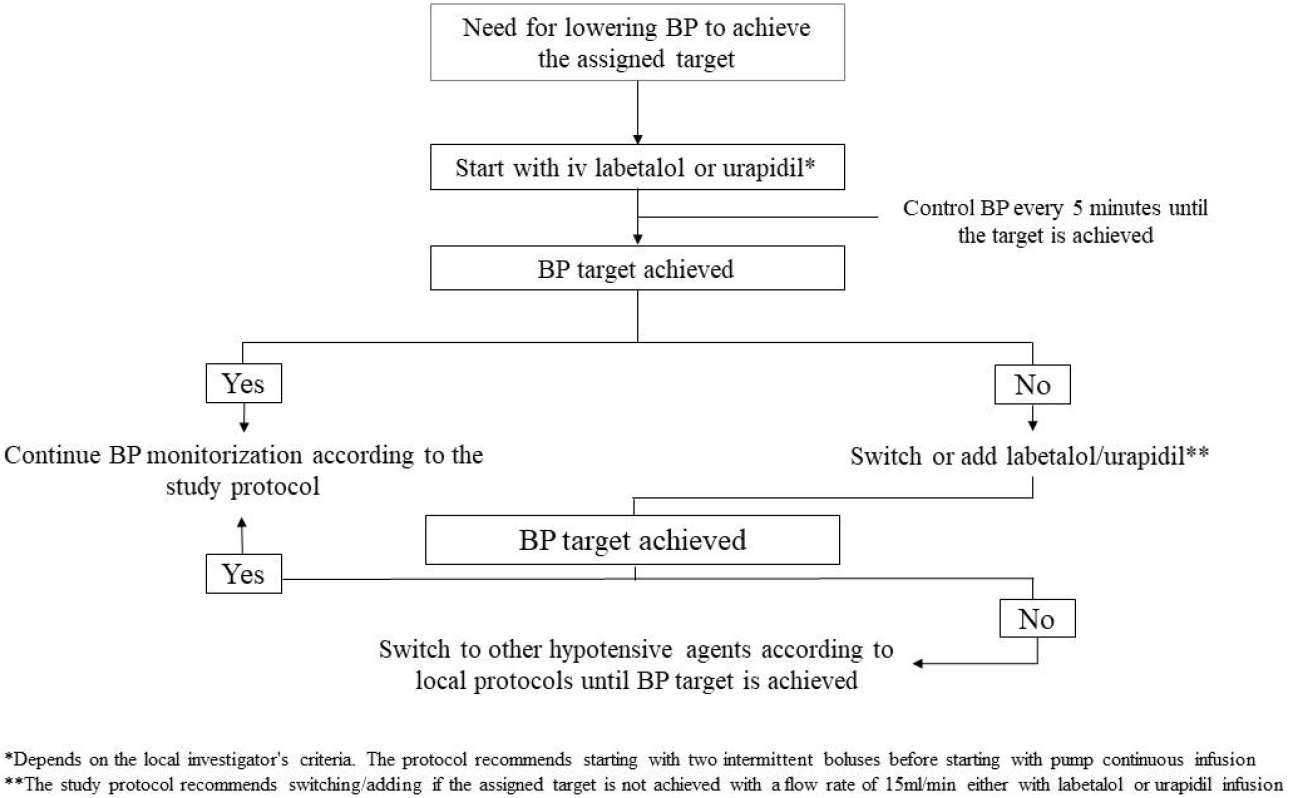
Study protocol for lowering blood pressure

**Fig 3:**
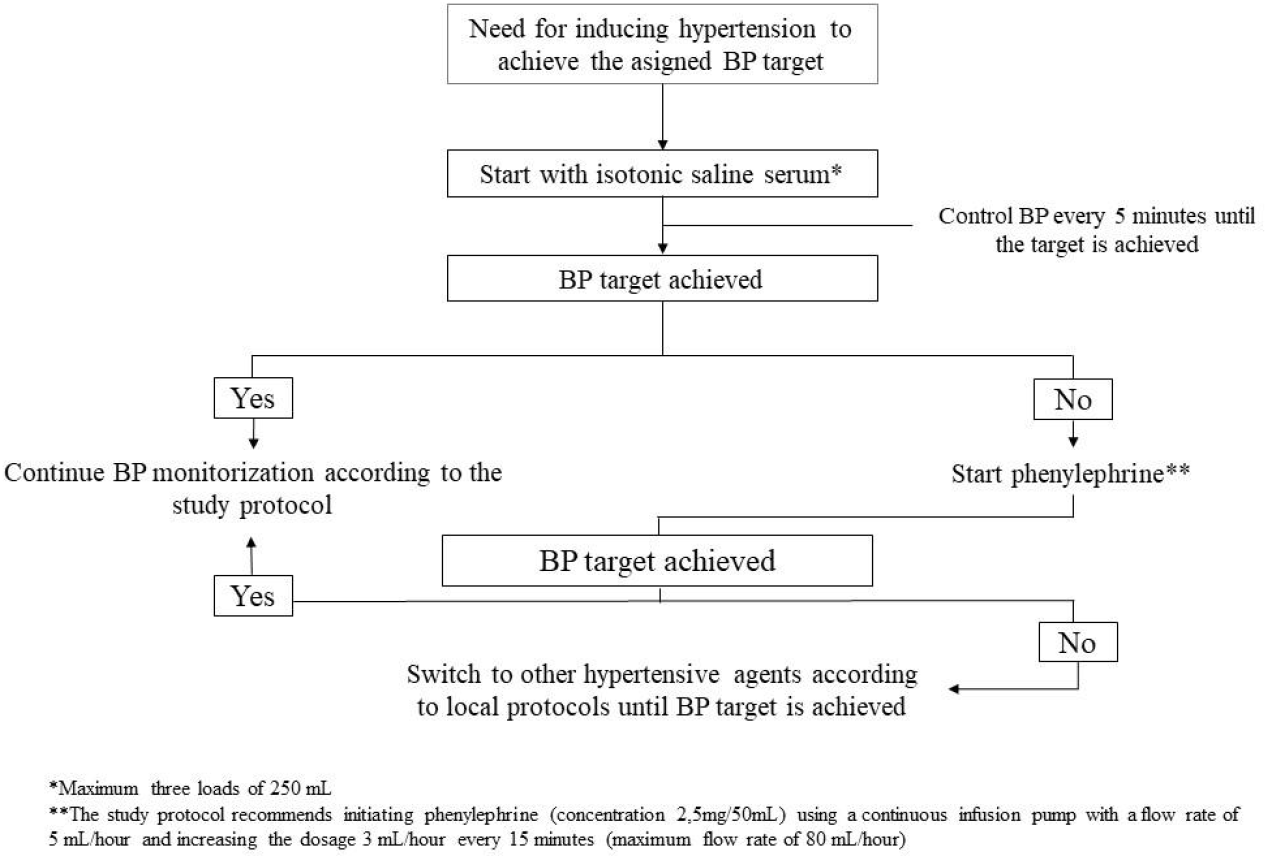
Study protocol for vasopressor therapy

The study protocol was interrupted promptly if a hemorrhagic transformation parenchymal hemorrhage type 2 (PH2) and/or any symptomatic ICH (sICH) according to the Heidelberg classification was detected during the first 72 h after EVT, if the patient experienced an early stroke recurrence or the patient presented with hemodynamic instability.

### Study Outcomes

#### Primary efficacy outcome

The primary outcome of the trial will be the proportion of favorable functional outcome at 90 days in an intention-to-treat analysis (ITT), in which all patients randomly assigned to one of the arms will be analyzed together, regardless of whether they completed or received the assigned intervention (ITT set). This outcome will be measured using the whole range of the mRS, which assesses daily functioning through the categorization of levels of disability. The mRS scaling is: 0 = no symptoms at all; 1 = no significant disability despite symptoms, but able to carry out all usual duties and activities; 2 = slight disability, unable to carry out all previous activities but able to look after own affairs without assistance; 3 = moderate disability requiring some help, but able to walk without assistance; 4 = moderate-severe disability, unable to walk without assistance and unable to attend to own bodily needs without assistance; 5 = severe disability, bedridden and requiring constant nursing care and attention; 6 = dead. Scores of 0-2 will be defined as a favorable outcome. Contrariwise, scores 3-6 will be defined as unfavourable outcomes. The mRS is evaluated locally at each recruiting centre by certified personnel blinded to the trial intervention.

#### Key secondary efficacy outcomes

The secondary efficacy outcomes of the trial will include (1) the shift on the mRS score at 90 days, also in the ITT set; (2) the proportion of favorable functional outcome (mRS 0-2) in the per-protocol (PP) set and (3) the shift on the mRS score at 90 days, also in the PP set.

#### Key safety outcomes

The secondary safety outcomes of the trial will include (1) neurological deterioration within 72 h (defined as a worsening of the neurological deficit ≥4 on the NIHSS); (2) sICH within 72 h; (3) any PH2; and (4) mortality at 90 days.

#### Other secondary outcomes

Other secondary outcomes will comprise the following: (1) hemorrhagic transformation of any type on brain imaging at 72 hours of treatment as defined by the Heidelberg bleeding classification [7]; (2) final infarct volume at the 24h CT scan; (3) final infarct volume at the 72h CT scan; (4) excellent functional outcome defined as an mRS score of 0-1; (5) Delta NIHSS defined as the difference between NIHSS at 24h and baseline NIHSS and; (6) Delta NIHSS between NIHSS at 72h and baseline NIHSS.

Radiological outcomes (hemorrhagic transformation and final infarct volume) will be assessed centrally by a Core Lab.

#### Protocol violations

Patients who have one or more of the following protocol violations will be excluded from the per-protocol (PP) population: age <18 years; prior mRS score >2; baseline ASPECTS<6; not achieving the assigned BP target within one hour from randomization; a time-in-target range (TTR) <40% during the study intervention and failure to obtain a blind assessment of the 90-day outcome.

The protocol deviations will be documented and reported as the number of subjects with a deviation. A comprehensive list of all protocol deviations will be given, which will provide insight into the extent and nature of deviations from the study protocol.

#### Loss to follow-up

Missing outcome data due to loss to follow-up will be handled using the Last Observation Carried Forward (LOCF) method, whereby the last available observation for each participant will be carried forward to impute missing values.

#### CRF, Data Safety and Monitoring Board and interim analyses

All data are entered into an electronic Case Record Form (CRF) and monitored by external qualified staff. The electronic CRF is managed according to general and specific standards of Good Clinical Practice. All serious adverse events are reviewed by a study monitor. An independent Data Safety and Monitoring Board (DSMB) has been in charge of monitoring the safety and the general development of the trial. The study protocol considered two formal interim analyses of 250 and 500 subjects for efficacy and futility. An additional interim analysis was performed after the publication of ENCHANTED 2/MT trial to discard harm in the already randomized population (n=130 at that moment). In each interim analysis, overwhelming efficacy/harm was estimated using O’Brien–Fleming boundaries on the binary outcome of 90-day mRS 0–2 versus 3–6. The criterion for determination of futility was based on the conditional power, defined as the probability of rejecting the null hypothesis at the final analysis given the data accumulated and under the assumption that the alternative hypothesis under the original design was true. A fall below 20% on the conditional power was considered a criterion of futility. After these interim analyses, the DSMB recommended continuing with the inclusion without changes in the protocol.

#### Sample Size Estimates

We originally based the sample size estimation on prior observational data from Goyal et al [8]. In this study, the investigators reported a proportion of patients with favorable outcomes (mRS 0–2) at 3 months of follow-up among patients with successful recanalization of 50% in the group with permissive BP management (<185/105 mm Hg) compared to 70% in the group with a BP target <160/90 mm Hg during the first 24 h (p = 0.041).

Considering these prior findings, accepting an alpha risk of 0.05 and a beta risk of 0.2 in a two-sided test, we calculated that 100 subjects would be necessary for the control group (routine BP management) and 100 in the intervention group (hemodynamic optimization) to find as statistically significant a proportion difference of 20%. Due to lack of randomized data at the time of designing the trial, and the fact that the arms of the HOPE trial were not directly comparable to the groups established by Goyal M et al., we finally decided to establish a minimum proportion difference of 10% (half of the difference found by Goyal M et al.) to assure that the trial would have enough power to conclude in favor or against the null hypothesis. Therefore, we established that 407 subjects would be necessary for the control group (routine BP management) and 407 in the intervention group (hemodynamic optimization). It was anticipated a drop-out rate of 5%.

After the publication of the meta-analysis of the first four clinical trials of BP management following EVT [9], we recalculated the sample size. We took as a reference the difference reported to be statistically significant between groups in the meta-analysis, which was 10%. The difference was the same as our initial estimation, although the proportion of favorable outcomes in the reference group was slightly higher. According to this new estimation of the sample size, accepting an alpha risk of 0.05 and a beta risk of 0.2 in a two-sided test, 393 subjects were necessary in each arm. For both sample size estimations, we used the normal corrected method using Stata v17 (TX, USA).

## 4. Statistical Analysis

### Description of the study population

Baseline characteristics of the patients will be presented by the treatment group in Table 1. Categorical variables will be presented as counts and percentages. Percentages will be calculated according to the number of patients in whom data are available. If missing values are important, the denominator will be added in a footnote in the corresponding table. Continuous variables will be summarised by use of standard measures of central tendency and dispersion, either mean and standard deviation (SD) for variables with a normal distribution according to the Shapiro-Wilk test, or median and interquartile range (IQR) if not normal.

For comparisons between groups, the chi-square test will be used for categorical variables, the independent t test or the Mann–Whitney U test (Wilcoxon rank-sum test) for continuous variables (depending on normality), and generalized estimating equations (GEE) for repeated measures. A p value < 0.05 will be considered statistically significant.

### Blood pressure control in both treatment arms

The target of SBP stratified according to final mTICI represents the main trial intervention. SBP control and variability will be summarized also by treatment groups and by final mTICI (2b or 2c/3) in Table 2. The following parameters of tendency and dispersion will be described:

- Mean of SBP and SD in each treatment arm
- Variability of SPB as assessed by the coefficient of variation (CV).
- Variability of SBP as assessed by the Average Real Variability (ARV).
- Time-in-target ratio (TTR) in each treatment group
- Use of vasopressors according to the study protocol summarized by the number of patients requiring vasopressors during the study period divided by the total number of patients in the corresponding arm.
- Use of hypotensive agents according to the study protocol summarized by the number of patients requiring BP lowering during the study period divided by the total number of patients in the corresponding arm.
- Frequency of serious hypotension drops defined as episodes of SBP <100mmHg in each treatment arms.

Tendency of SBP will be also displayed by treatment groups in Figure 2 by depicting serial error bars at 6-hour intervals.

### Primary outcome analysis

A direct comparison between the two trial arms will be made considering the score on the mRS at 90 (+/-15) days after randomization. This will be an ITT analysis. The primary effect parameter will be the unadjusted odds ratio of achieving functional independence (mRS 0–2) with its 95% confidence interval using logistic regression. To increase the power of the study, a sensitivity analysis of the primary outcome with adjustment for the key prognostic covariates of age, baseline NIHSS, prior mRS score and time from onset to randomization will be performed.

### Secondary efficacy outcome analyses

The secondary efficacy outcome analyses will include:

- The unadjusted common odds ratio of a shift in the direction of a better outcome on the mRS with its 95% confidence interval. The common odds ratio will be estimated with ordinal logistic regression. This will also be an ITT analysis. Again, a sensitivity analysis of the primary outcome with adjustment for the key prognostic covariates of age, baseline NIHSS, prior mRS score and time from onset to randomization will be performed
- Sensitivity analyses using the PP set will be performed for the two prior efficacy outcomes: (1) dichotomic functional outcome (mRS 0-2 vs. 3-6) using binary logistic regression and (2) the shift analysis using ordinal logistic regression. How the PP set will be established is described before in “Protocol violations”.

### Secondary safety analyses

The following safety outcomes will be presented as the odds of presenting the adverse event comparing both arms with its 95% confidence interval. These odds ratios will be estimated using logistic regression analysis and will be adjusted with the same statistical approach as the effect analyses except for the prior mRS, that will not be included as an adjustment covariate. These safety outcomes include (1) neurological deterioration within 72 h; (2) sICH within the first 72 h; (3) any PH2; and (4) mortality at 90 days.

### Other secondary outcomes

No formal adjustments will be performed to constrain the overall type I error associated with the secondary exploratory analyses. All these secondary analyses will be performed in the ITT set. The preplanned analyses for these secondary outcomes will be as follows:

- Hemorrhagic transformation of any type: frequencies and percentages for each type of hemorrhagic transformation will be presented by the treatment group. A chi-square test will be used to evaluate the differences between both arms.
- Final infarct volume: FIV will be described as median mL with its IQR for each arm of the trial. A Mann Whitney U test will be used to assess differences between the optimized group vs. the control group.
- Excellent functional outcome (mRS score of 0-1): this variable will be presented also with frequencies and percentages according to the assigned group. Again, a chi-square test will be used to evaluate the differences between both arms.
- Delta NIHSS: this parameter will be calculated at two time points 24h and 72h. The difference in the NIHSS score will always take the baseline NIHSS as a reference. It will be presented as a median with its IQR for each arm of the trial. A Mann Whitney U test will be used to assess differences between the optimized group vs. the control group.

### Subgroup analyses

Subgroup analyses will be carried out irrespective of whether there is a significant treatment effect on the primary outcome. These analyses will be unadjusted and using the ITT set. They will be presented in Figure 3 as a forest plot with a p value for interaction for each category.

Analysis of 8 key subgroups will be carried out for the primary outcome.

- Age: <80 versus ≥80 years
- Sex: male versus female
  - Baseline NIHSS score categorized as:
  - NIHSS ≤10
  - NIHSS 11–15
  - NIHSS 16–20
  - NIHSS ≥21
- ASPECTS: 6-8 vs. 9-10
- Tandem occlusion vs. isolated TICA/M1/M2 occlusions
- Collateral status in baseline CTA (good vs. poor according to the TAN score)
- Final mTICI: 2b vs. 2c/3
- Time from stroke onset to admission <6h vs. extended time window (6-24h)

The primary method for analyzing each subgroup will be a logistic regression model incorporating an interaction test. To quantify the treatment-by-subgroup interaction, the change in the log likelihood will be determined when the interaction term is added to the logistic regression model, which already includes the main effects of treatment and subgroup.

### Tables and figures

The following tables will be presented:

- Table 1: collected baseline characteristics of the participants by treatment group.
- Table 2: SBP control and variability by treatment groups and by final mTICI (2b or 2c/3).
- Table 3: primary and secondary efficacy outcomes.
- Table 4: secondary safety outcomes.

In addition, the following figures will be prepared:

- Figure 1. : CONSORT diagram illustrating the flow of patients through the study.
- Figure 2. : Tendency of SBP in both treatment groups by serial box plots at 6-hour intervals.
- Figure 3. : mRS distribution at 90 days from randomization depicted in horizontal bars according to treatment groups.
- Figure 4. : Forest plot showing subgroup analyses for the primary outcome.

## Data Availability

All data produced in the present study are available upon reasonable request to the authors

## Abbreviation

ACA: Anterior cerebral artery
ASPECTS: Alberta Stroke Program Early CT Score
BA: Basilar artery
BP: Blood pressure
CI: Confidence interval
CRF: Case Report Form
CT: Computed Tomography
CTA: Computed Tomography Angiography
DSMB: Data Safety Monitoring Board
ECASS II: European Cooperative Acute Stroke Study II
EVT: Endovascular Treatment
HI: Hemorrhagic infarction
HT: Hemorrhagic Transformation
ICA: Internal carotid artery
ICH: Intracerebral hemorrhage
IQR: Interquartile range
ITT: Intention-to-treat
MCA: Middle cerebral artery
mRS: Modified Rankin Scale
mTICI: Modified Treatment In Cerebral Infarction
NIHSS: National Institute of Health Stroke Scale
OR: Odds ratio
PCA: Posterior cerebral artery
PH: Parenchymal hematoma
PP: Per-protocol
PROBE: Prospective, randomized, open-label trial with blinded end-point assessment
SBP: Systolic Blood Pressure
sICH: Symptomatic Intracranial Hemorrhage
TOAST: Trial of Org 10 172 in acute stroke treatment

## References

[1] Mazighi M, Richard S, Lapergue B, Sibon I, Gory B, Berge J, et al. Safety and efficacy of intensive blood pressure lowering after successful endovascular therapy in acute ischaemic stroke (BP-target): a Multicentre, open-label, Randomised Controlled Trial. Lancet Neurol. 2021;20:265–74.

[2] Yang P, Song L, Zhang Y, Zhang X, Chen X, Li Y, et al. Intensive blood pressure control after endovascular thrombectomy for acute ischaemic stroke (ENCHANTED2/MT): a Multicentre, open-label, blinded-endpoint, Randomised Controlled Trial. Lancet. 2022;400:1585–96.

[3] Nam HS, Kim YD, Heo JN, Lee H, Jung JW, Choi JK, et al. Intensive vs conventional blood pressure lowering after endovascular thrombectomy in acute ischemic stroke: the OPTIMAL-BP randomized clinical trial. JAMA. 2023;330:832–42.

[4] Mistry EA, Hart KW, Davis LT, Gao Y, Prestigiacomo CJ, Mittal S, et al. Blood pressure management after endovascular therapy for acute ischemic stroke: the BEST-II randomized clinical trial. JAMA. 2023;330:821–31.

[5] Powers WJ, Rabinstein AA, Ackerson T, Adeoye OM, Bambakidis NC, Becker K, et al. 2018 guidelines for the early management of patients with acute ischemic stroke: a guideline for Healthcare professionals from the American heart association/American stroke association. Stroke. 2018;49:e46–110.

[6] Camps-Renom P, Guasch-Jiménez M, Martínez-Domeño A, Prats-Sánchez L, Ramos-Pachón A, Álvarez-Cienfuegos J, et al. A Randomized Trial on Hemodynamic Optimization of Cerebral Perfusion after Successful Endovascular Therapy in Patients with Acute Ischemic Stroke (HOPE). Cerebrovasc Dis. 2025; 54:559–566.

[7] von Kummer R, Broderick JP, Campbell BCV, Demchuk A, Goyal M, Hill MD, et al. The Heidelberg bleeding classification: classification of bleeding events after ischemic stroke and reperfusion therapy. Stroke. 2015;46(10):2981–6.

[8] Goyal N, Tsivgoulis G, Pandhi A, Chang JJ, Dillard K, Ishfaq MF, et al. Blood pressure levels post mechanical thrombectomy and outcomes in large vessel occlusion strokes. Neurology. 2017;89:540–7.

[9] Gharaibeh K, Aladamat N, Mierzwa AT, Rao R, Alhajala H, Al Kasab S, et al. Blood Pressure after Successful Endovascular Therapy: A Systematic Review and Meta-Analysis of Randomized Control Trials. Ann Neurol. 2024;95:858–865.

